# Accuracy of the diagnostic tests for the detection of Chagas disease: a systematic review and meta-analysis

**DOI:** 10.1101/2022.09.03.22279561

**Authors:** Mayron Antonio Candia-Puma, Laura Yesenia Machaca Luque, Brychs Milagros Roque Pumahuanca, Alexsandro Sobreira Galdino, Rodolfo Cordeiro Giunchetti, Eduardo Antonio Ferraz Coelho, Miguel Angel Chávez-Fumagalli

**Affiliations:** Computational Biology and Chemistry Research Group, Vicerrectorado de Investigación, Universidad Católica de Santa María, Arequipa 04000, Peru; Facultad de Ciencias Farmacéuticas, Bioquímicas y Biotecnológicas, Universidad Católica de Santa María, Arequipa 04000, Peru; Laboratório de Biotecnologia de Microrganismos, Universidade Federal São João Del-Rei, Divinópolis, Minas Gerais, Brazil; Laboratório de Biologia das Interações Celulares, Instituto de Ciências Biológicas, Universidade Federal de Minas Gerais, Minas Gerais, Brazil; Instituto Nacional de Ciência e Tecnologia em Doenças Tropicais, INCT-DT, Salvador 40015-970, Bahia, Brazil; Programa de Pós-Graduação em Ciências da Saúde: Infectologia e Medicina Tropical, Faculdade de Medicina, Universidade Federal de Minas Gerais, Belo Horizonte, Minas Gerais, Brazil; Departamento de Patologia Clínica, COLTEC, Universidade Federal de Minas Gerais, Belo Horizonte, Minas Gerais, Brazil

**Keywords:** Chagas disease, Diagnostic Tests, Meta-analysis, Systematic review, Sensitivity and Specificity

## Abstract

The present systematic review and meta-analysis about the accuracy of diagnostic tests aim to describe the findings of literature over the last thirty years for the diagnosis of Chagas disease (CD). This work aimed to determine the accuracy of diagnostic techniques for CD in the disease’s acute and chronic phases. The PubMed database was searched for studies published between 1990 and 2021 on CD diagnostic. Fifty-six published studies that met the criteria were analyzed and included in the meta-analysis, evaluating diagnostic accuracy through sensitivity and specificity. For Enzyme-Linked Immunosorbent Assay (ELISA), Fluorescent Antibody Technique (IFAT), Hemagglutination Test (HmT), Polymerase Chain Reaction (PCR) and (Real-Time Polymerase Chain Reaction qPCR diagnosis methods, the sensitivity had a median of 99.0%, 78.0%, 75.0%, 76.0% and 94.0%, respectively; while specificity presented a median of 99.0%, 99.0%, 99.0%, 98.0% and 98.0%, respectively. This meta-analysis showed that ELISA and qPCR techniques had a higher performance compared to other methods of diagnosing CD in the chronic and acute phases, respectively. It was concluded by means of the AUC restricted to the false positive rates, that the ELISA diagnostic test presents the highest performance in diagnosing acute and chronic CD, compared to serological and molecular tests. Future studies focusing on new CD diagnostics approaches should be targeted.

## Introduction

Chagas disease (CD) is an anthropozoonosis caused by the protozoan parasite *Trypanosoma cruzi*, which is transmitted mainly by blood-sucking bugs (also known as “kissing-bug”) from the subfamily *Triatominiae* [1,2] Other transmission forms are vertical transmission from mother to child or by blood transfusion, organ transplant, laboratory accident, oral contamination, and breastfeeding [3]. Over six million people are affected by the disease in Latin America, being endemic in 21 countries [4–6]. Additionally, it has been proposed that in the United States approximately 300,000 persons live with the infection, including 57,000 Chagas cardiomyopathy patients and 43,000 infected reproductive-age women [7], even though only a small fraction are properly diagnosed and treated [8]. Comparably, in the last decade, globalization has allowed the disease to spread through European countries, such as Austria, Belgium, France, Germany, Italy, Netherlands, Portugal, Spain, Sweden, Switzerland, the United Kingdom, Australia, Japan, and Canada [9,10]. In this scenario, at least 100 million people have a high risk of infection by living in endemic areas of disease [11], while the estimated annual global burden is $627.46 million in healthcare costs and 800,000 disability-adjusted life-years [7,15], besides that, approximately 10,000 deaths per year can be attributed to the disease [16], making CD a serious public health problem worldwide.

The infection has two distinct clinical phases separated by an indeterminate period [12] In the acute phase, the disease is characterized by high parasitemia; usually asymptomatic or oligosymptomatic; and patients can exhibit fever, anorexia, tachycardia, and cutaneous manifestations, such as Chagoma’s and Romaña’s signs [13].; In the chronic phase, the infection can manifest as neurological, cardiac, digestive, and cardio-digestive alterations [14]. CD cardiomyopathy is the most severe and life-threatening manifestation of the disease; affecting about 40% of patients in the chronic phase and appearing as heart failure, arrhythmia, heart block, thromboembolism, stroke, and sudden death [15].; Mega-visceral syndromes are caused by denervation of the enteric nervous system that appears years after the acute infection and includes Megacolon (the commonest form), Megaesophagus, and Chagasic enteropathy, where about 10% of asymptomatic patients with chronic CD have radiological gastrointestinal abnormalities [16]; as well, the neurological manifestations of CD manifests as neuritis, which results in distorted tendon reflexes and sensory impairment, while is reported in up to 10% of the patients. Isolated central nervous system involvement cases can also include dementia, confusion, chronic encephalopathy, and sensitive and motor deficits [17].

Control strategies applied to CD combine two approaches, which include the prevention, diagnosis, and treatment of infected individuals [18]; however, despite being a disease that has been discovered more than a century ago, there is little access to diagnosis and treatment in primary health care [19,20]. Benznidazole (BNZ) and Nifurtimox (NFX) are employed currently drugs in the therapy against CD, while BNZ treatment is counter-indicated for pregnant women and people with significant hepatic and renal illness; NFX is recommended as a second-line drug, only in the cases where BNZ failures and in absence of neurological and psychiatric disorders [21]; likewise, both are not effective in the chronic phase of the disease and produce severe adverse reactions [22]. Otherwise, potential vaccine candidates against CD have been investigated in recent decades; however, none have passed the preclinical stages of development. [23,24].

Laboratory diagnostic tests for CD depend largely on the clinical stage of the disease, while in the acute phase, it allows the direct detection of the parasite using molecular biology (polymerase chain reaction and real-time polymerase chain reaction) or parasitological (xenodiagnoses) techniques; oppositely, in the chronic phase, parasitemia becomes low and intermittent [25–28]; still, acute infection leads to seroconversion and anti-*T. cruzi*-specific immunoglobulins are detectable for years, so the infection can be indirectly identified by serological methods, such as enzyme-linked immunosorbent assay (ELISA), complement fixation test (CFT), immunofluorescent antibody technique (IFAT), hemagglutination test (HmT), radioimmunoprecipitation assay (RIPA), and western blot (WB) [29]. However, at present, there is no gold standard diagnostic test, since commercial tests have shown a high rate of false-positive results [30], for this reason, the World Health Organization (WHO) recommends that the diagnosis of CD should be carried out using two conventional tests based on the detection of different antigens [31]; and in the case of ambiguous or discordant results, a third technique should be used [32]. This situation reveals the urgent need for the development of new diagnostic tools for disease diagnosis [34–36]. A satisfactory method will allow the establishment of a patient registry with CD, a useful tool to provide information on its epidemiology, characteristics, and treatment [33]. Additionally, it must be considered that a behavioral design that allows establishing the reasons for people’s refusal to participate in diagnostic campaigns for this disease can alter the internal and external validity of the diagnosis [34].

The objective of the current work is to systematically review and summarize the available literature on the diagnostic accuracy of diagnostic tests for CD. For this, a systematic review of the medical literature was carried out over the period from 1990 to 2021. Results were analyzed through a meta-analysis based on the techniques used in diagnosing CD. The diagnostic techniques examined were PCR, qPCR, Xenodiagnosis, ELISA, CFT, IFAT, HmT, RIPA, and WB. Thus, we hope that the data generated will help to identify the basic need to fund the research organization for the screening, improvement, and effectiveness of CD diagnosis.

## Results

### Search results and characteristics of the selected studies

In the current work, a systematic review followed by a meta-analysis to measure the accuracy of diagnostic tests for CD was performed, and a flowchart of the strategy employed to select the studies is shown (Figure 1). For this, a search with the MeSH Terms “*Chagas Disease*” was performed in the Pubmed database, followed by the construction of a network map of the co-occurrence of MeSH terms; the search resulted in 370 published papers in a 1990–2021-year range, whereas establishing the value of five as the minimum number of occurrences of keywords, a map with 969 keywords that reaches the threshold was constructed (Figure 2A). In the analysis of the map, it is shown that five main clusters were formed, while terms such as “*Enzyme-linked immunosorbent assay*”, “*Polymerase chain reaction*”, “*Real-Time Polymerase Chain Reaction*”, “*Xenodiagnosis*”, “*Complement Fixation Tests*”, “*Fluorescent Antibody Technique*”, “*Hemagglutination Tests*”, “*Radioimmunoprecipitation Assay*” and “*Blotting, Western*”, associated with diagnostic techniques were observed in the fifth cluster (purple color). Also, terms such as “*Chagas disease*”, “*humans*”, “*Trypanosoma cruzi”*, “*female*”, “*trypanocidal agents*”, “*adult*”, and “*insect vectors*” were recurrent denominators (Figure 2A).

**Figure 1.**
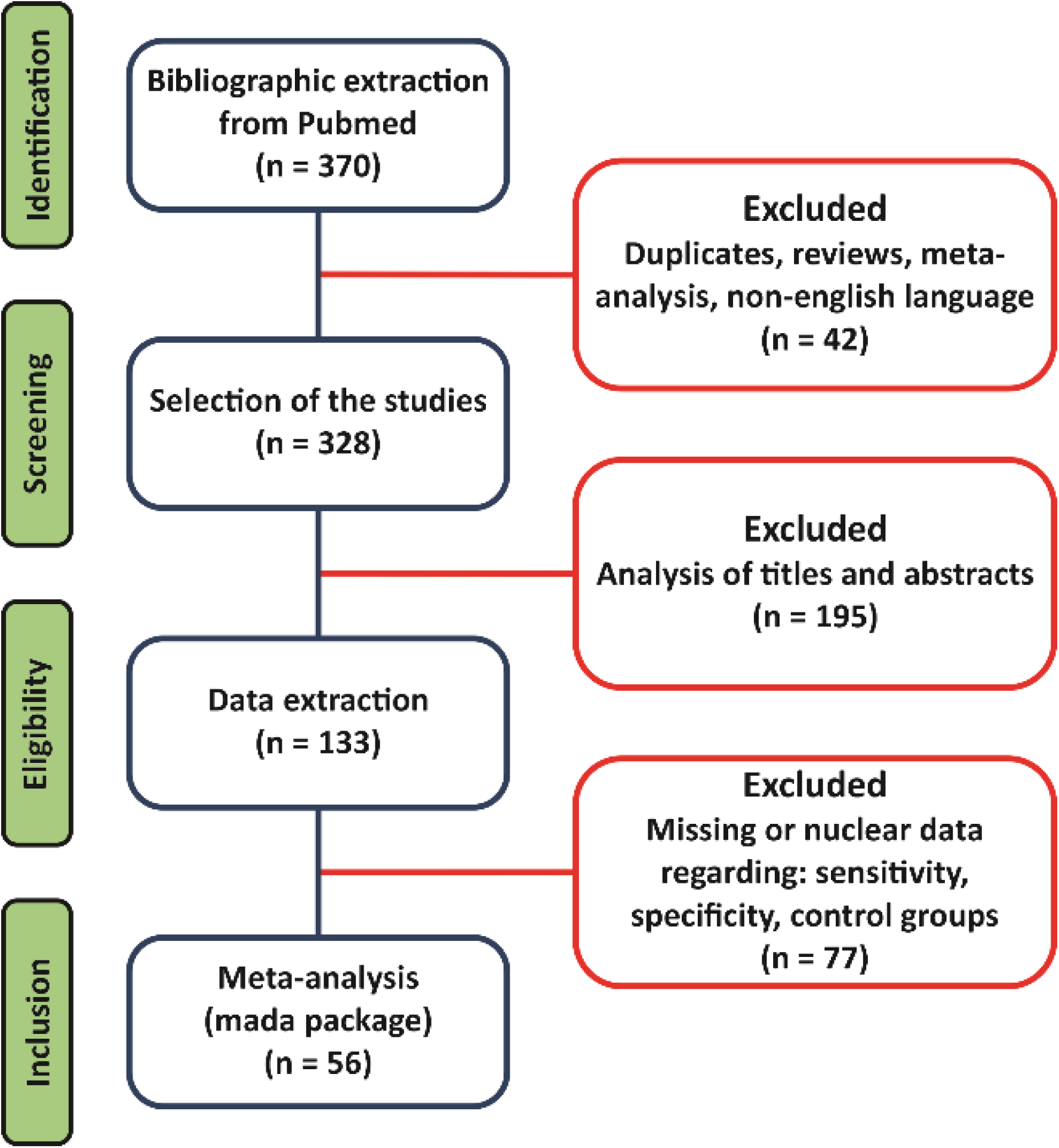
Systematic review and meta-analysis workflow diagram.

**Figure 2.**
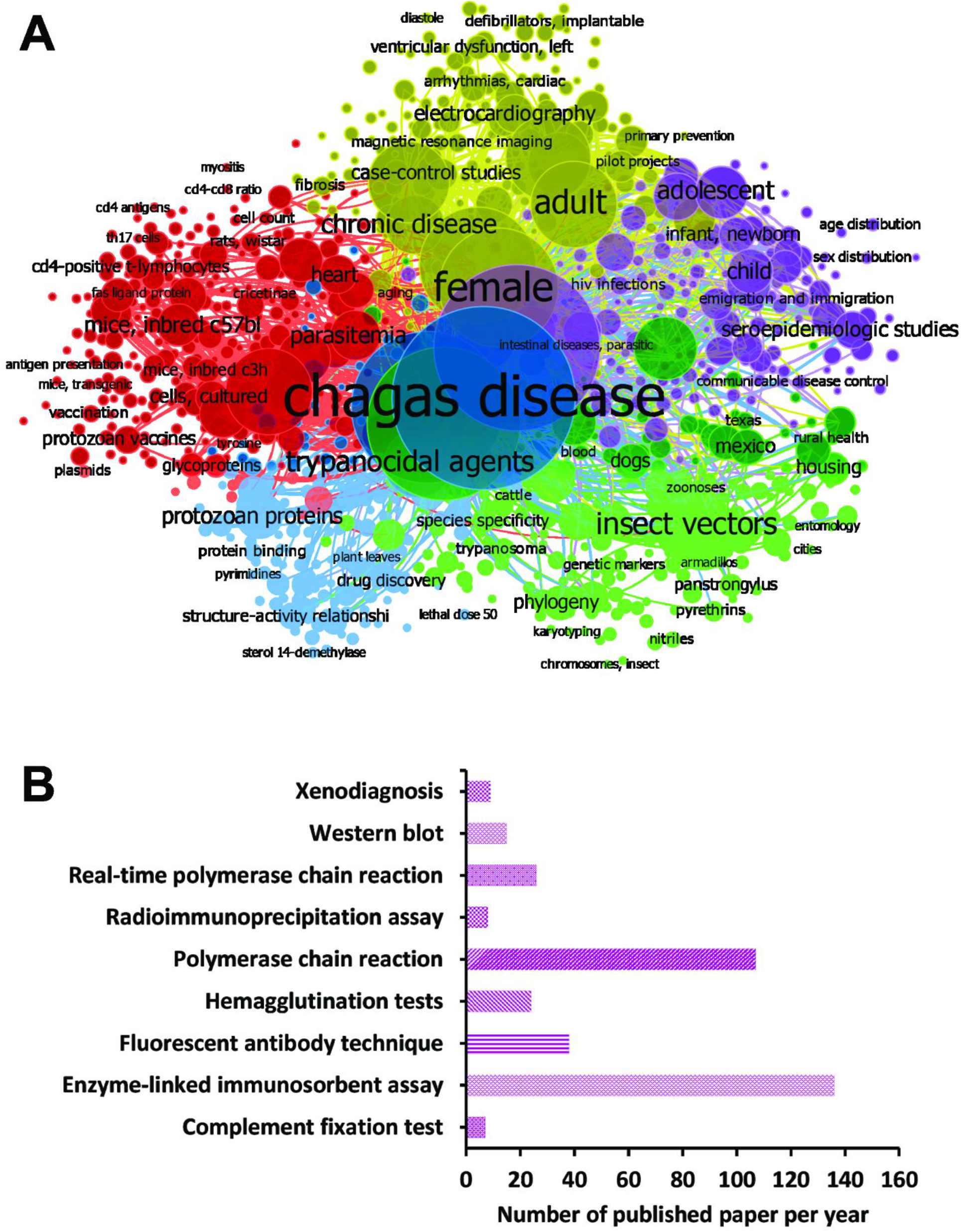
Papers were selected for the different diagnostic techniques using MeSH terms in the PubMed database. A) Bibliometric map created by VOSviewer based on MeSH terms co-occurrence. B) Number of papers found in the search for each diagnostic technique found in cluster analysis.

The terms identified in this first analysis were employed in a second search at the Pubmed database, while associated each new term with the terms “*Chagas Disease*” and “*Sensitivity and Specificity*”; forming the new search strings: “*Chagas Disease*” [MeSH Terms] AND “*Sensitivity and Specificity*” [MeSH Terms] AND “*Polymerase Chain Reaction*” [MeSH Terms] for PCR; “*Chagas Disease*” [MeSH Terms] AND “*Sensitivity and Specificity*” [MeSH Terms] AND “*Real-Time Polymerase Chain Reaction* “ [MeSH Terms] for qPCR; “*Chagas Disease*” [MeSH Terms] AND “*Sensitivity and Specificity*” [MeSH Terms] AND “*Xenodiagnosis*” [MeSH Terms] for XD; “*Chagas Disease*” [MeSH Terms] AND “*Sensitivity and Specificity*” [MeSH Terms] AND “*Enzyme-Linked Immunosorbent Assay* “ [MeSH Terms] for ELISA; “*Chagas Disease*” [MeSH Terms] AND “*Sensitivity and Specificity*” [MeSH Terms] AND “*Complement Fixation Tests*” [MeSH Terms] for CFT; “*Chagas Disease*” [MeSH Terms] AND “*Sensitivity and Specificity*” [MeSH Terms] AND “*Fluorescent Antibody Technique* “ [MeSH Terms] for FAT; “*Chagas Disease*” [MeSH Terms] AND “*Sensitivity and Specificity*” [MeSH Terms] AND “*Hemagglutination Tests*” [MeSH Terms] for HmT; “*Chagas Disease*” [MeSH Terms] AND “*Sensitivity and Specificity*” [MeSH Terms] AND “*Radioimmunoprecipitation Assay*” [MeSH Terms] for RIPA; and “*Chagas Disease*” [MeSH Terms] AND “*Sensitivity and Specificity*” [MeSH Terms] AND “*Blotting, Western*” [MeSH Terms] for WB.

Besides, the number of selected studies for PCR, qPCR, XD, ELISA, CFT, FAT, HmT, RIPA, and WB was: 107, 26, 9, 136, 7, 38, 24, 8, and 15, respectively (Figure 2B). Though using a three-step eligibility criterion, 237 articles were excluded in the first two steps. In the last stage of data extraction, 41 articles were excluded. Therefore, 92 articles were selected for meta-analysis. Also, data on geographical characteristics extracted from the 92 selected articles were analyzed (Figure 3); while in most of the studies, the diagnostic technique employed was ELISA (Figure 3A); additionally, the American continent has carried out a greater number of studies, with Brazil being the country where a higher number of the population of Chagasic patients, in general, have been studied (Figure 3B, C, and D).

**Figure 3.**
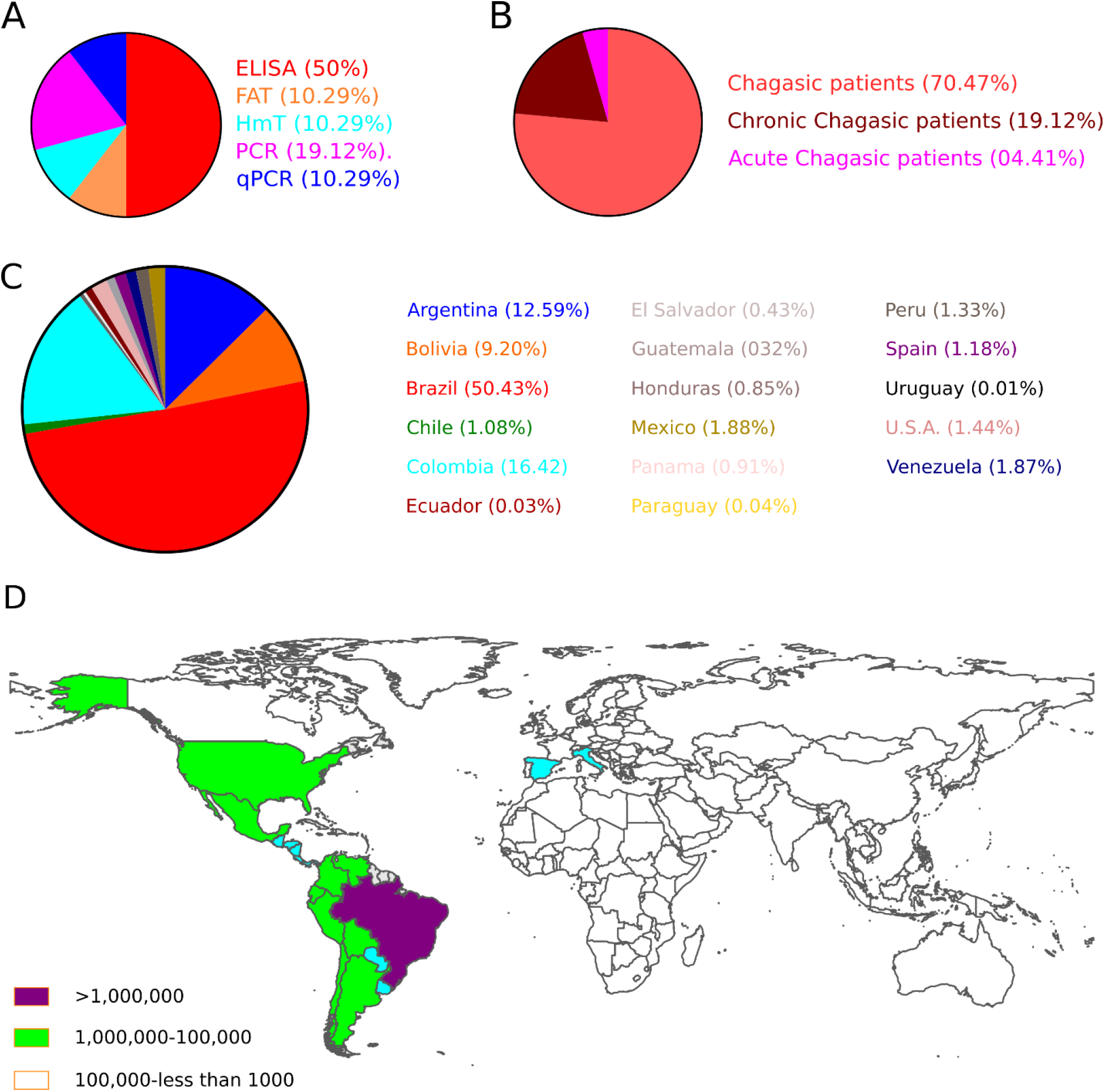
The geographical location of Chagas’s disease studies. A) The pie chart shows the biomarkers; B) the clinical description of patients; and C) the Number of CD patients included in the selected studies worldwide. D) Estimative of the global prevalence of Chagas disease, 2017 [8]

### Meta-analysis of the diagnostic techniques for CD

#### Enzyme-linked immunosorbent assay

Thirty-four studies were selected for the ELISA technique: [35–68] in which a total of 6,054 subjects were studied. Sensitivity ranged from 78.0% to 100%, with a median of 99.0%, CI 95% (94, 100), while the test for equality of sensitivities presented a χ^2^ = 657.24, df = 33, p-value = 2e-16. Study specificity ranged from 83.0 to 100%, with a median of 99.0%, 95%CI (95, 100); the test for equality of specificities showed χ^2^ = 311.4699, df = 33, p-value = 2e-16. Additionally, the results regarding LR+ {median 71.74, 95% CI (13.71, 476.73)}, LR-{median 0.01, 95%CI (0.00, 0.08)} and DOR {median 5938.93, 95%CI (331.76, 100154.55)}. The analyzed diagnostic performance is summarized in Fig. 4 and Supplementary Fig. 1.

**Figure 4.**
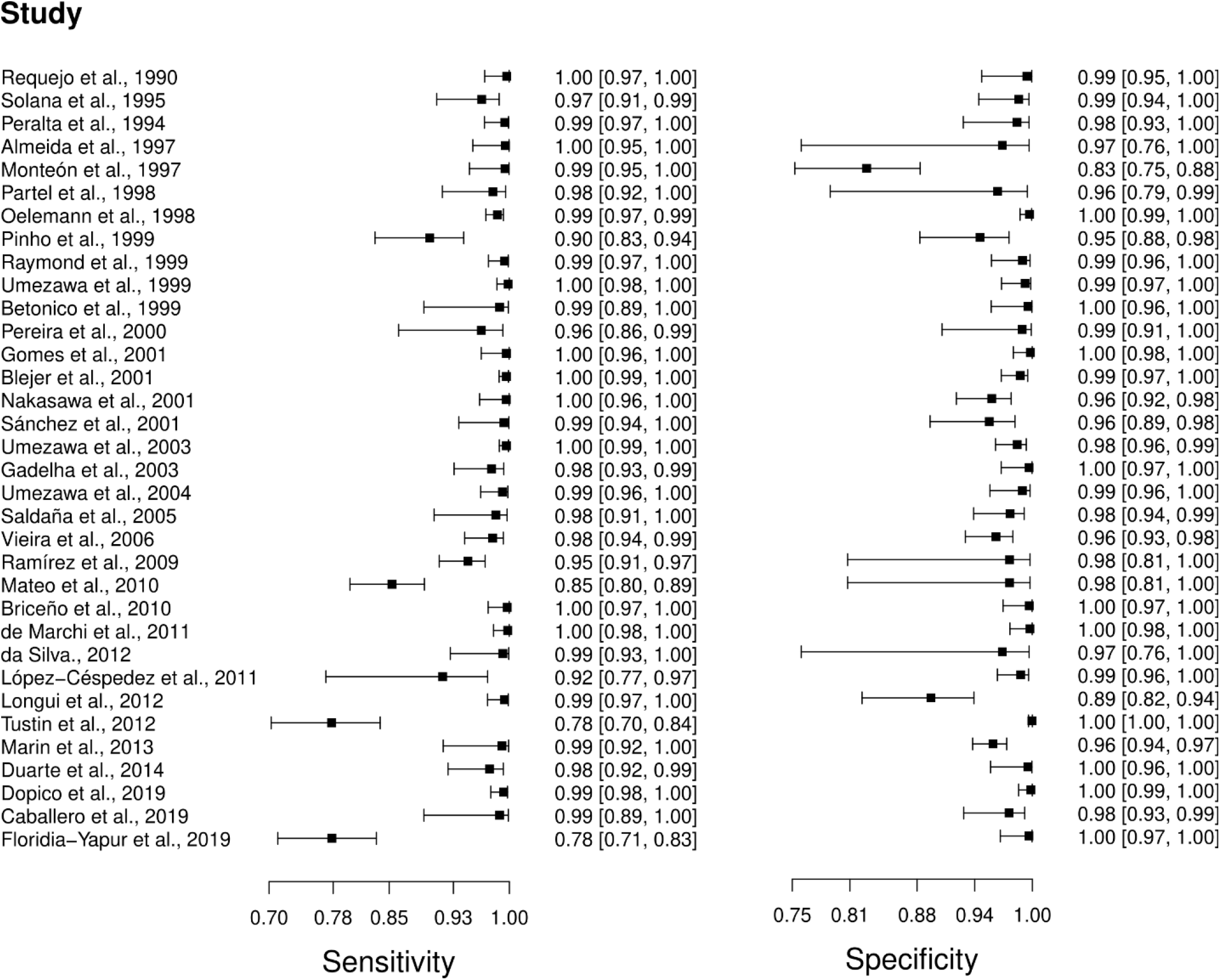
Study data and paired forest plot of the sensitivity and specificity of Enzyme-linked immunosorbent assay (ELISA) in Chagas’s disease diagnosis. Data from each study are summarized. Sensitivity and specificity are reported with a mean (95% confidence limits). The Forest plot depicts the estimated sensitivity and specificity (black squares) and its 95% confidence limits (horizontal black line).

#### Fluorescent Antibody

Seven studies were selected for the FAT diagnostic technique: (56,57,77-81). A total of 810 subjects were studied. Sensitivity ranged from 74.0 to 78.0%, with a median of 78.0%, 95% CI (70, 84); while the test for equality of sensitivities showed: χ^2^ = 0.064, df =6, p-value =1. The specificity of the studies ranged from 92.0 to 100%, with a median of 99.0%, 95% CI (89, 100); while the test for equality of specificities presented χ^2^ = 7.89, df = 6, p-value = 0.246. In addition, the results regarding LR+ {median 62.22, 95%CI (3.95, 979.16)}, LR-{median 0.23, 95%CI (0.16, 0.31)} and DOR {median 62.22, 95%CI (3.95, 979.16)} are displayed. The analyzed diagnostic performances are summarized in Fig. 5 and Supplementary Fig. 2.

**Figure 5.**
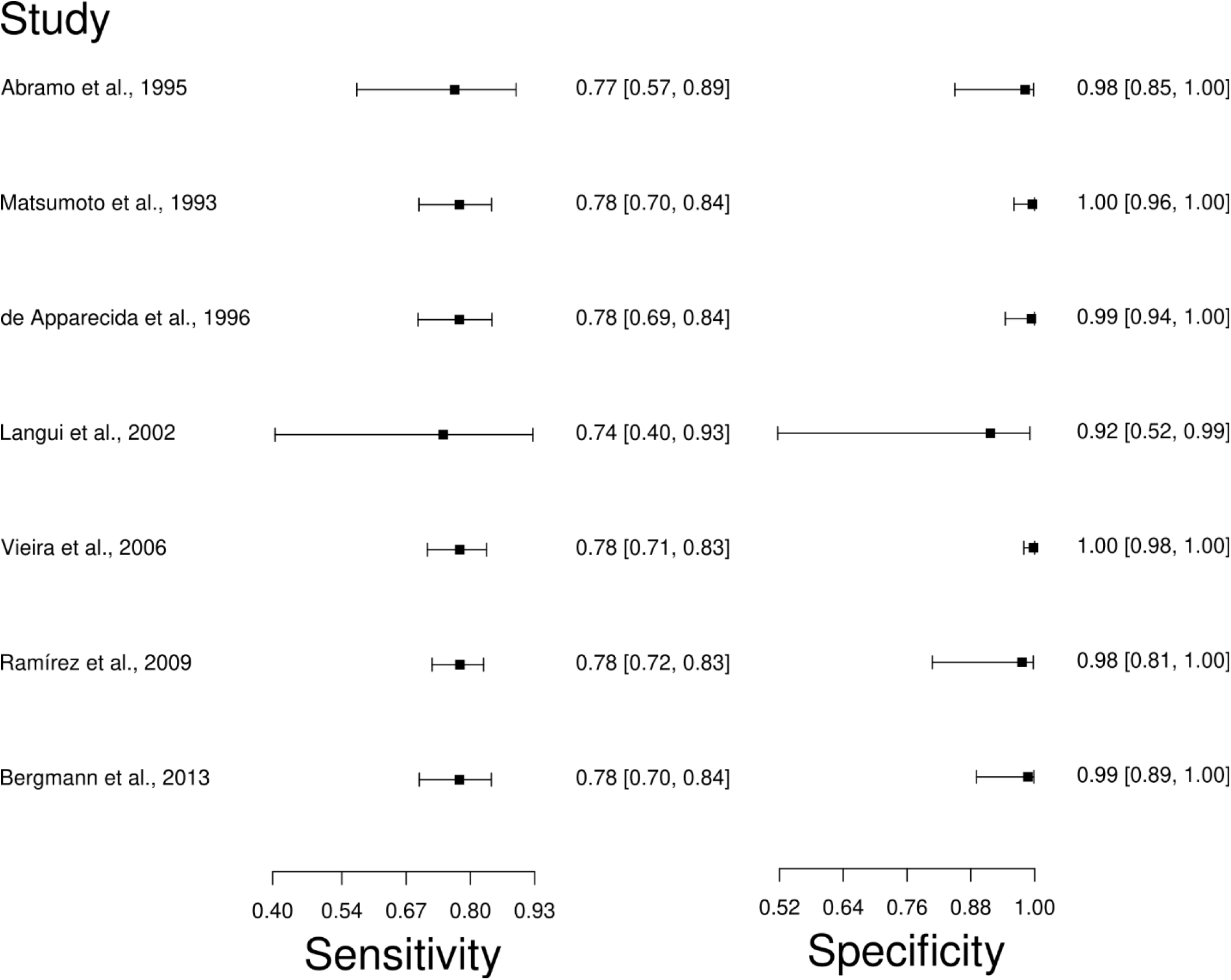
Study data and paired forest plot of the sensitivity and specificity of Fluorescence antibody assay (FAT) in Chagas’s disease diagnosis. Data from each study are summarized. Sensitivity and specificity are reported with a mean (95% confidence limits). The Forest plot depicts the estimated sensitivity and specificity (black squares) and its 95% confidence limits (horizontal black line).

#### Hemagglutination Tests

The analysis identified 24 published studies that used HmT as a diagnostic technique for CD. After analysis, only 7 studies [52,55,65,69–72] were selected. A total of 1450 subjects were studied. The sensitivity of the studies ranged from 53.0 to 99.0%, with a median of 75.0%, and 95%CI (70, 80). Test for equality of sensitivities analysis showed: χ^2^ = 162.98, df = 6, p-value = 2e-16. The specificity of the studies ranged from 98.0 to 100%, with a median of 99.0%, 95%CI (96, 100); while the test for equality of specificities: χ^2^ = 5.19, df = 6, p-value = 0.51. In addition, the results regarding LR+ {median 76.42, 95%CI (15.72, 1201.06)}, LR-{median 0.25, 95%CI (0.20, 0.30)} and DOR {median 633.85, 95%CI (76.19, 5332.02)} are displayed. The diagnostic performance of the selected studies is summarized in Fig. 6 and Supplementary Figure 3.

**Figure 6.**
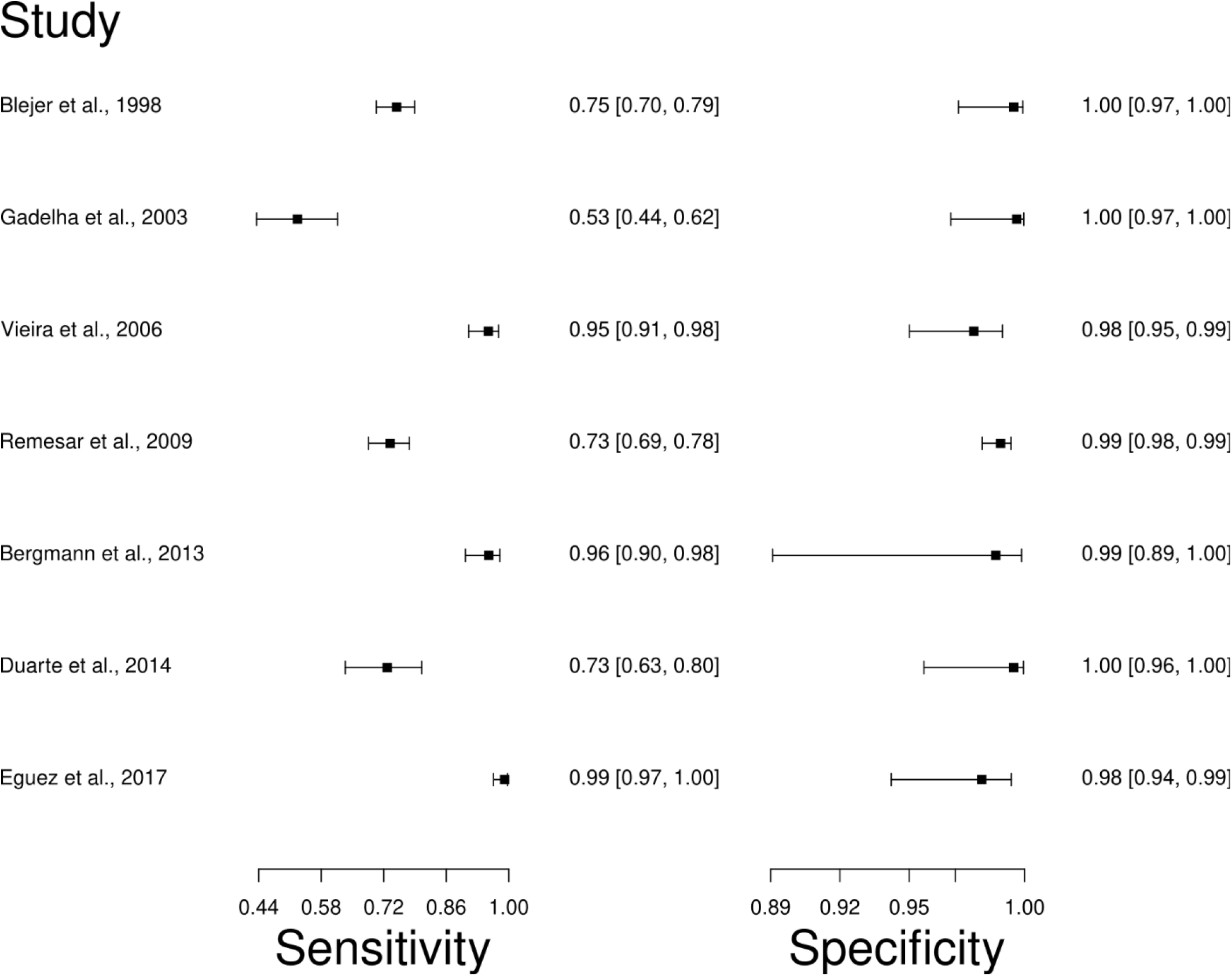
Study data and paired forest plot of the sensitivity and specificity Hemagglutination test (HmT) in Chagas’s disease diagnosis. Data from each study are summarized. Sensitivity and specificity are reported with a mean (95% confidence limits). The Forest plot depicts the estimated sensitivity and specificity (black squares) and its 95% confidence limits (horizontal black line).

#### Polymerase chain reaction

Thirteen studies were selected for the PCR diagnostic technique [55,56,65,73–82]. A total of 2198 subjects were studied. Sensitivity ranged from 2.0 to 99%, with a median of 76.0%, CI 95% (67, 84), while the test for equality of sensitivities presented a χ^2^ = 516.43, df = 12, p-value = 2e-16. Study specificity ranged from 45.0 to 100%, with a median of 98.0%, 95%CI (82, 100); the test for equality of specificities showed χ^2^ = 315.74, df = 12, p-value = 2e-16. In addition, the results regarding LR+ {median 18.32, 95% CI (2.59, 251.81)}, LR-{median 0.25, 95%CI (0.17, 0.36)} and DOR {median 163, 95%CI (9.1, 2920.82)}. The analyzed diagnostic performance is summarized in Fig. 7 and Supplementary Fig. 4.

**Figure 7.**
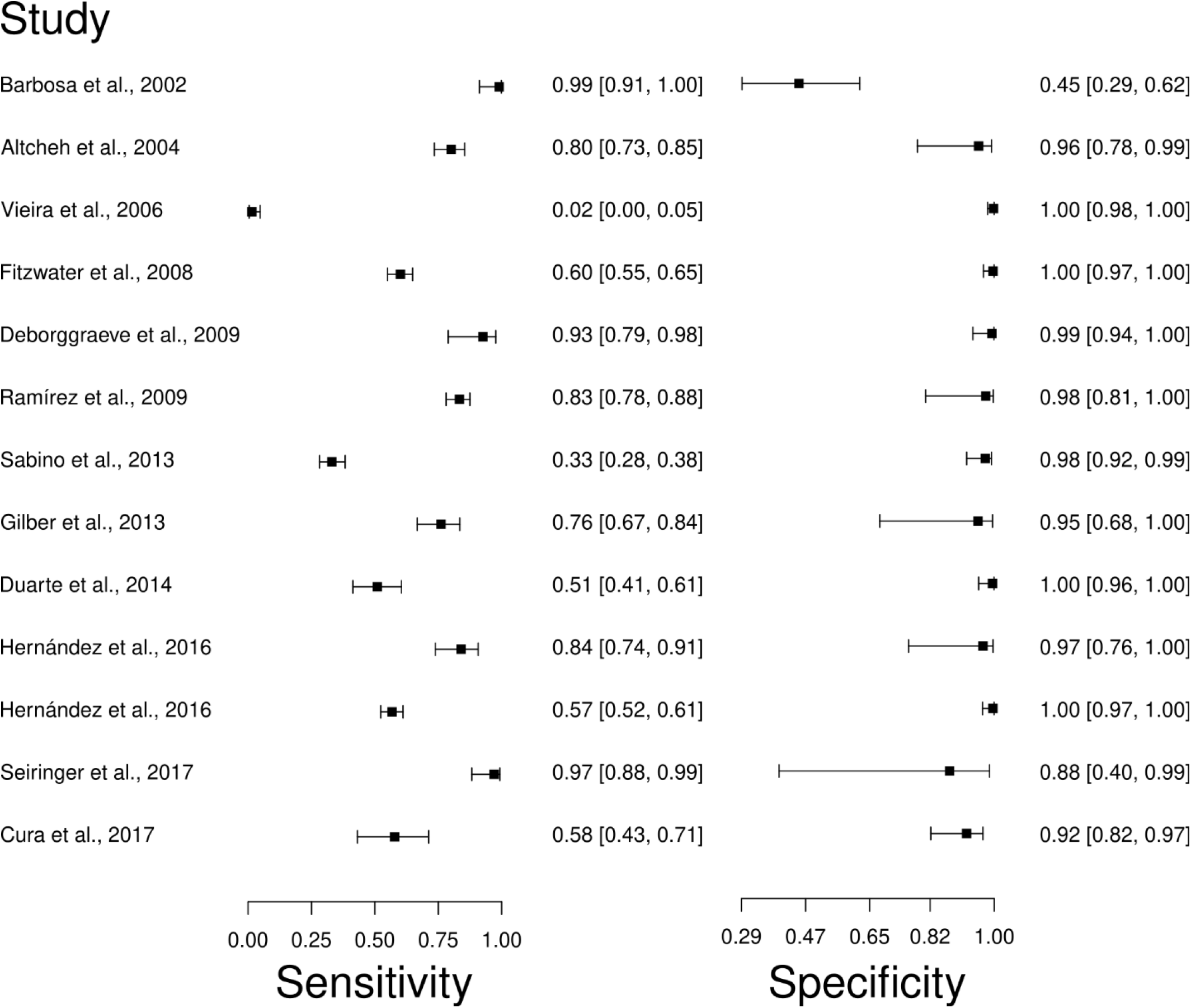
Study data and paired forest plot of the sensitivity and specificity Polymerase chain reaction (PCR) in Chagas’s disease diagnosis. Data from each study are summarized. Sensitivity and specificity are reported with a mean (95% confidence limits). The Forest plot depicts the estimated sensitivity and specificity (black circles) and its 95% confidence limits (horizontal black line).

#### Real-time polymerase chain reaction

The analysis identified 26 published studies that used qPCR as a diagnostic technique for CD. After analysis, only 7 studies [79,80,82–86] were selected. A total of 995 subjects were studied. Sensitivity ranged from 40.0 to 100%, with a median of 94.0%, 95% CI (82, 98); while the test for equality of sensitivities showed: χ^2^ = 122.39, df =6, p-value =2e-16. The specificity of the studies ranged from 79.0 to 100%, with a median of 98.0%, 95% CI (83, 100); while the test for equality of specificities presented χ^2^ = 81.46, df = 6, p-value = 1.78e-15. In addition, the results regarding LR+ {median 46.92, 95%CI (3.01, 730.43)}, LR-{median 0.06, 95%CI (0.02, 0.21)} and DOR {median 597.2, 95%CI (31.35, 12131.7)} are displayed. The analyzed diagnostic performances are summarized in Fig. 8 and Supplementary Fig. 5.

**Figure 8.**
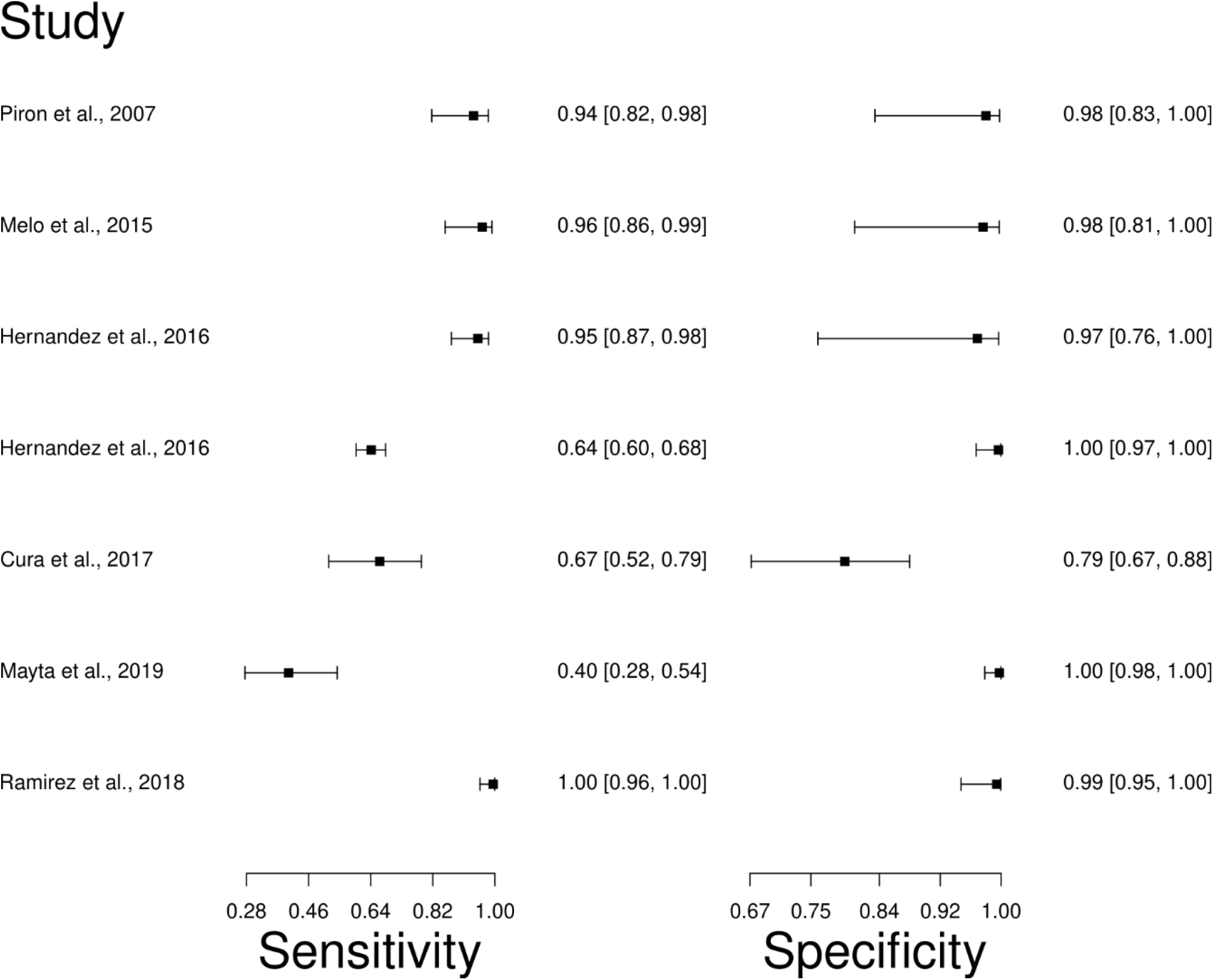
Study data and paired forest plot of the sensitivity and specificity quantitative Polymerase chain reaction (qPCR) in Chagas’s disease diagnosis. Data from each study are summarized. Sensitivity and specificity are reported with a mean (95% confidence limits). The Forest plot depicts the estimated sensitivity and specificity (black squares) and its 95% confidence limits (horizontal black line).

#### Other techniques

Regarding the XD, CFT, RIPA, and WB diagnostic techniques, one [87], one [88], three [89–91], and three [92–94] studies were selected, respectively. While according to the criteria established in the workflow no analysis was performed regarding these diagnostic techniques.

#### Summary ROC curves (sROC)

sROC curve analysis was conducted to compare diagnostic data from ELISA, FAT, HmT, PCR, and qPCR techniques for chronic CD (Figure 9), due to differences in sensitivity and specificity, which can be generated by implicit or explicit variations across studies and variation in test cut-off points. The area under the curve (AUC) calculated for ELISA, FAT, HmT, PCR, and qPCR was 0.989, 0.770, 0.988, 0.957, and 0.981, respectively, indicating slightly better performance for the ELISA in chronic CD. Likewise, when the AUC was restricted to the observed false positive rates (FPR) (AUC_FPR_) the results showed a relatively better performance of the ELISA diagnostic test for chronic CD (Figure 9). Also, data from ELISA and qPCR for acute CD were compared by sROC curve analysis. The AUC calculated for ELISA and qPCR was 0.986 and 0.987, respectively, indicating the similar performance of both techniques. However, when the AUC_FPR_ was calculated, the results showed a better performance of the ELISA diagnostic test for acute CD (Figure 10).

**Figure 9.**
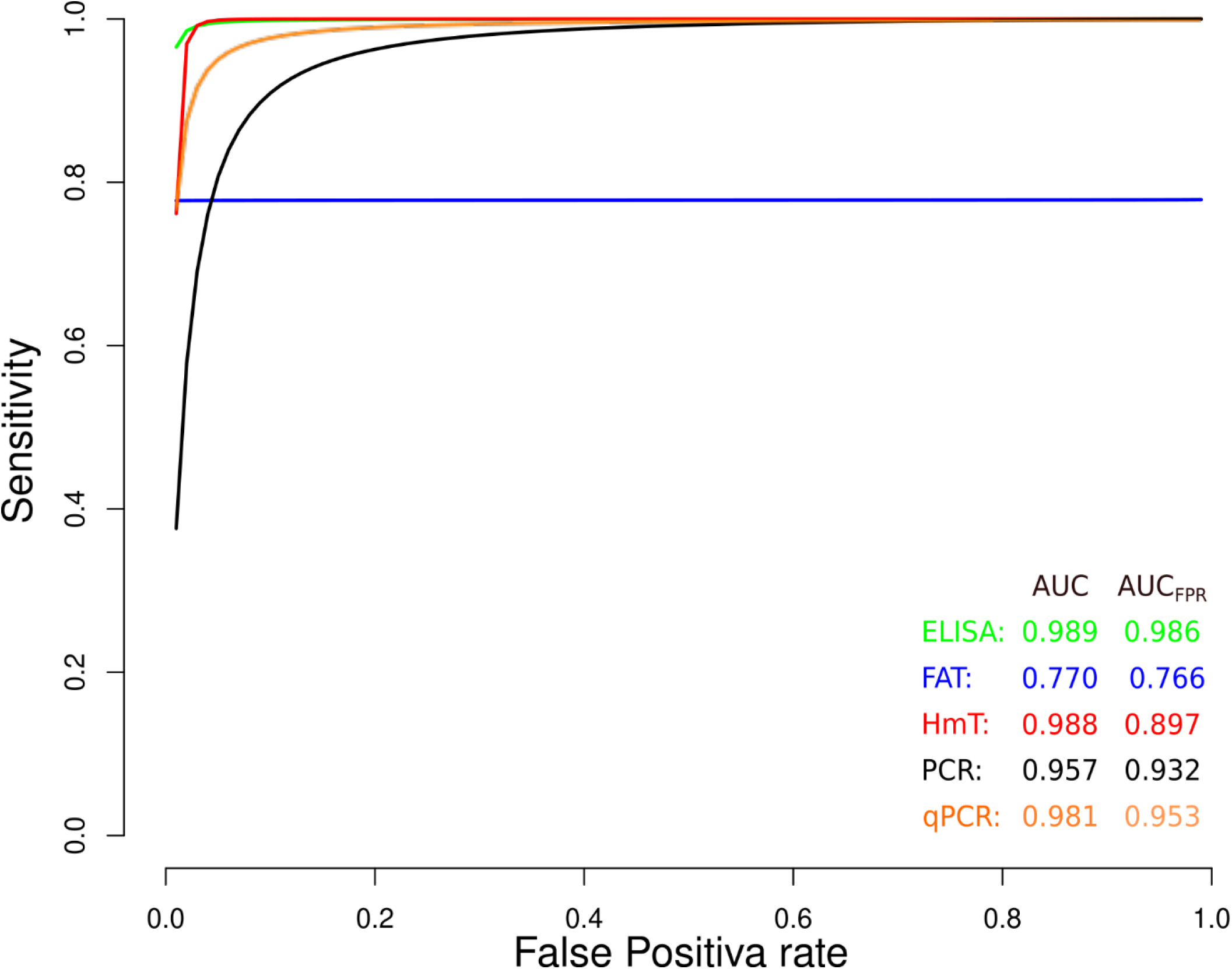
Meta-analysis of diagnostic test accuracy analysis. Summary receiver operating curve (sROC) plot of false positive rate and sensitivity. Comparison between ELISA, FAT, HmT, PCR, and qPCR methods in the diagnosis of chronic Chagas disease

**Figure 10.**
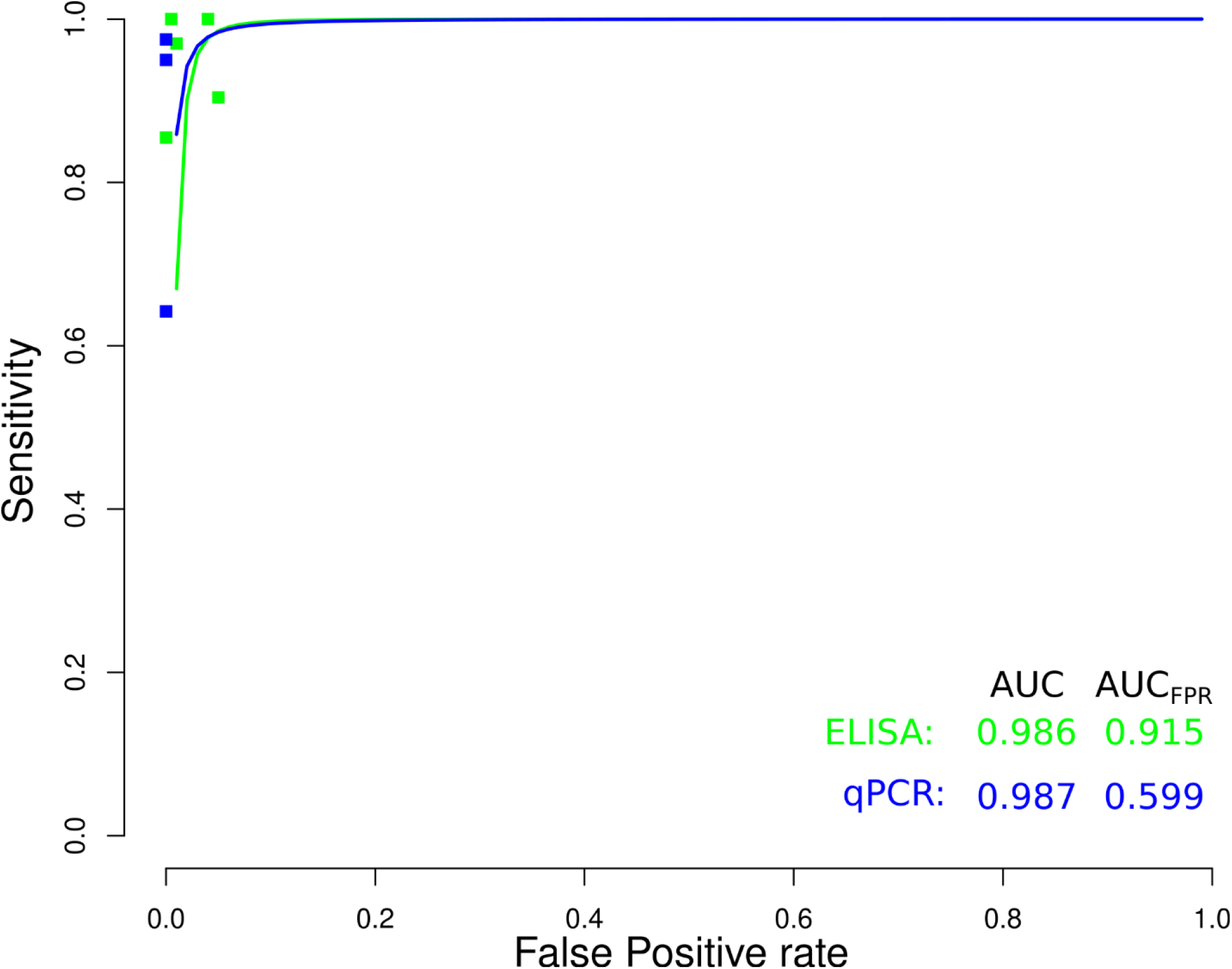
Meta-analysis of diagnostic test accuracy analysis. Summary receiver operating curve (sROC) plot of false positive rate and sensitivity. Comparison between ELISA and qPCR methods in the diagnosis of acute Chagas disease.

## Discussion

CD was once confined to rural areas in Latin America, where it was commonly transmitted through vectors [95], while in recent years; increased migration has been pointed out as the main driver of the urbanization of the disease, which is a broad and complex response to changing labor needs and agricultural, demographic, and geopolitical conditions [96]. Therefore, the migratory flow has been key to the appearance of CD in areas where it was not previously reported, which makes CD a global concern that expands its geographical location in an increasingly globalized world [97]. In addition, it is estimated that 90% of people with CD are unaware of their infection and are therefore at risk of transmitting the disease and suffering from complications [98], which is aggravated since the dynamics of parasitemia during infection fluctuate [99]. Furthermore, diagnostic techniques employed in diagnosing CD have insufficient accessibility, sensitivity, specificity, and applicability, and most of them require expensive resources and equipment that are often unavailable in endemic areas [100–102]. Additionally, it has been proposed that the sensitivity and specificity of current tests are lower than generally reported in quality and in unblinded studies, leading to the potential for underdiagnosis [103].

In recent times, molecular methods, such as PCR and qPCR, have been introduced as supportive diagnostic tests for CD [107], these methods represent a great advance in DNA and/or RNA quantification in biological samples and have been extensively studied in the assessment of the parasitic load in patients [108,109].; However, crucial knowledge, such as the sampling methods, which have not been clearly stated; and the accuracy of these techniques for diagnosing CD in the different clinical phases, remains poorly understood [110]. Besides that, the analysis presented in the current work of qPCR data shows the best performance among molecular diagnostic methods, with a median sensitivity and specificity of 94.0 and 98.0%, respectively. It has been stated that the diagnostic efficacy of molecular techniques is high in the acute CD, while in the chronic phase the immunological techniques are more effective [111]; when comparing the ELISA to qPCR methods in the diagnosis of the acute CD results, it was shown no significant difference; but, when comparing the AUC restricted to the observed false positive rates, qPCR showed inferior results; which can be explained partially by the difference in the number of studies evaluated and sample sizes.

ELISA, HmT, and FAT are standard techniques applied to detect anti-*T. cruzi* antibodies, whereas these diagnostic methods have different antibody recognition rationales [99]; however, ELISA exhibits several advantages over other techniques because of its simplicity, selectivity, and sensitivity [112]; besides, the Pan-American Health Organization (PAHO) recommendations and other diagnostic guidelines advise the use of two different serological techniques for chronic Chagas disease diagnosis, one of the techniques being ELISA [113]; for these reasons, several ELISA-based diagnostic tests have been developed and approved by the United States Food and Drug Administration (FDA) for blood donor screening and clinical diagnostic testing, [114]. Regarding the results obtained in the present work, the ELISA method presented the best performance among immunological methods with a median sensitivity and specificity of 99.0 and 99.0%, respectively. Furthermore, when compared to other diagnosis methods accessed in the work, ELISA showed the best results in the diagnosis of acute and chronic CD, when the AUC was restricted to the observed false positive rates.

Although XD allows CD to be diagnosed at the subclinical stage of the disease, where there are no clinical signs [104]; and that serological tests, such as CFT, RIPA, and WB are preferably used for the diagnosis of CD during the chronic phase [105]; unexpectedly, the number of studies selected made it impossible to include them in the meta-analysis, which requires at least 5 studies for analysis with a *p* < 0.05 [106]. Yet, a search of single MeSH terms for “Chagas Disease”, “Sensitivity and Specificity”, “Xenodiagnosis”, “Complement Fixation Test”, “Radioimmunoprecipitation assay” and “Blotting, Western” showed 14 253, 633 656, 117, 16 780, 566, 163 653 studies, respectively, while combining them only 15, 7, 8 and 15 were found, correspondingly. Inherent flaws associated with a systematic review and meta-analysis studies, such: as the location and selection of studies, loss of information on important outcomes, inappropriate subgroup analyses, conflict with new experimental data, and duplication of publication [115], should be considered as limitations of the present work. Also, across the studies analyzed, one of the main problems found was the heterogeneity of groups studied, clinical settings, and diagnostic performance measurements, whereas biased estimates of sensitivity and specificity, which could tend to inflate estimates, were also common problems found in the analyzed studies.

## Conclusions

The accurate and sensitive diagnosis of CD is important for effective treatment and for the adoption of control measures against the disease. This study found that the ELISA technique showed better diagnostic performance in the chronic and acute phases of CD, when compared to other serological (HmT and FAT) and molecular (PCR and qPCR) techniques; suggesting then its feasibility to be used for the sensitive and specie CD diagnosis.

## Methods

### Search strategy

The systematic review search strategy employed to evaluate the literature was developed as described [106]. Briefly, Medical Subject Headings (MeSH) terms, which is a controlled vocabulary for indexing and searching biomedical literature [116], were employed in a search carried out at the PubMed database (https://pubmed.ncbi.nlm.nih.gov/), (last searched on 05 July 2021). Pubmed is the main database for the health sciences, generated by the National Center for Biotechnology Information (NCBI) at the National Library of Medicine (NLM), the database contains about 32 million citations, belonging to more than 5,300 journals currently indexed in MEDLINE [117]. While firstly, to find terms associated in the literature with CD diagnosis, the MeSH term “Chagas Disease” was employed alone, as the results were plotted into a network map of co-occurrence of MeSH terms in the VOSviewer software (version 1.6.17) [118], which employs a modularity-based method algorithm to measure the strength of clusters [119]. The resulting cluster content was analyzed to select relevant terms associated with diagnostic techniques. Lastly, a second search was designed with each MeSH term obtained in the cluster analysis, associated with the MeSH term *“sensitivity and specificity”*, which are commonly regarded as summary points of diagnostic accuracy of a test [120], and the MeSH term *“Chagas disease”*.

### Study selection and Data extraction

The studies were selected in three stages. In the first, non-English language articles, duplicate articles, reviews, and meta-analyses were excluded, only articles published after 1990 and conducted on humans were included. In the second stage, the titles and abstracts of the articles selected through the search strategy were examined. Finally, the highly relevant full studies were retrieved and separated from the articles with a title or abstract that did not provide sufficient data to be included. The information consigned for each study chosen included the diagnostic technique, the number, type, and clinical characteristics of patients with CD and healthy controls. All studies evaluating the sensitivity and specificity of CD diagnostic techniques have been included. Furthermore, the data related to the geographical distribution, the number of studies carried out by country, and the frequency of the diagnostic techniques used was extracted. Studies with unclear or missing data regarding the CD and healthy control groups or data describing the sensitivity and specificity obtained in the studies were excluded from further analysis.

### Statistical analysis

Results were entered into Microsoft Excel (version 10.0, Microsoft Corporation, Redmond, WA, USA) spreadsheets and analyzed in the R programming environment (version 4.0.3) using the package “*mada*” (version 0.5.10) (https://cran.r-project.org/web/packages/mada/index.html); which employs a hierarchical model that accounts for within and between-study (heterogeneity) and the correlation between sensitivity and specificity [121]. Initially, the number of true negatives (TP), false negatives (FN), true positives (TP), and false positives (FP) were analyzed separately for each diagnostic technique; while the evaluation of sensitivity (Se) and specificity (Sp) made possible to determine the diagnostic performance. Additionally, the positive likelihood ratio (LR +), the negative likelihood ratio (LR-), the diagnostic likelihood ratio (DOR), and the 95% confidence interval (CI) were determined. Summary receiver operating characteristics (sROC) curves were fitted, according to the parameters of the “*Reitsma*” model of the “*mada*” package, and were used to compare the diagnostic accuracy of CD diagnostic techniques [122]. The confidence level for all calculations was set to 95%, using a continuity correction of 0.5 if pertinent.

## Data Availability

Not applicable.

## Author Contributions

Conceptualization: M.A.C.-P., and M.A.C.-F.; data curation: M.A.C.-P., L.Y.M.L., B.M.R.P., and M.A.C.-F.; formal analysis: M.A.C.-P., and M.A.C.-F.; funding acquisition: M.A.C.-F.; investigation: A.S.G., R.C.G., and E.A.F.C.; methodology: M.A.C.-P., and M.A.C.-F.; writing—review & editing: A.S.G., R.C.G., E.A.F.C., and M.A.C.-F. All authors have read and agreed to the published version of the manuscript.

## Institutional Review Board Statement

Not applicable.

## Informed Consent Statement

Not applicable.

## Data Availability Statement

Not applicable.

## Acknowledgments

Not applicable.

## Figure Legends

**Supplementary 11.**
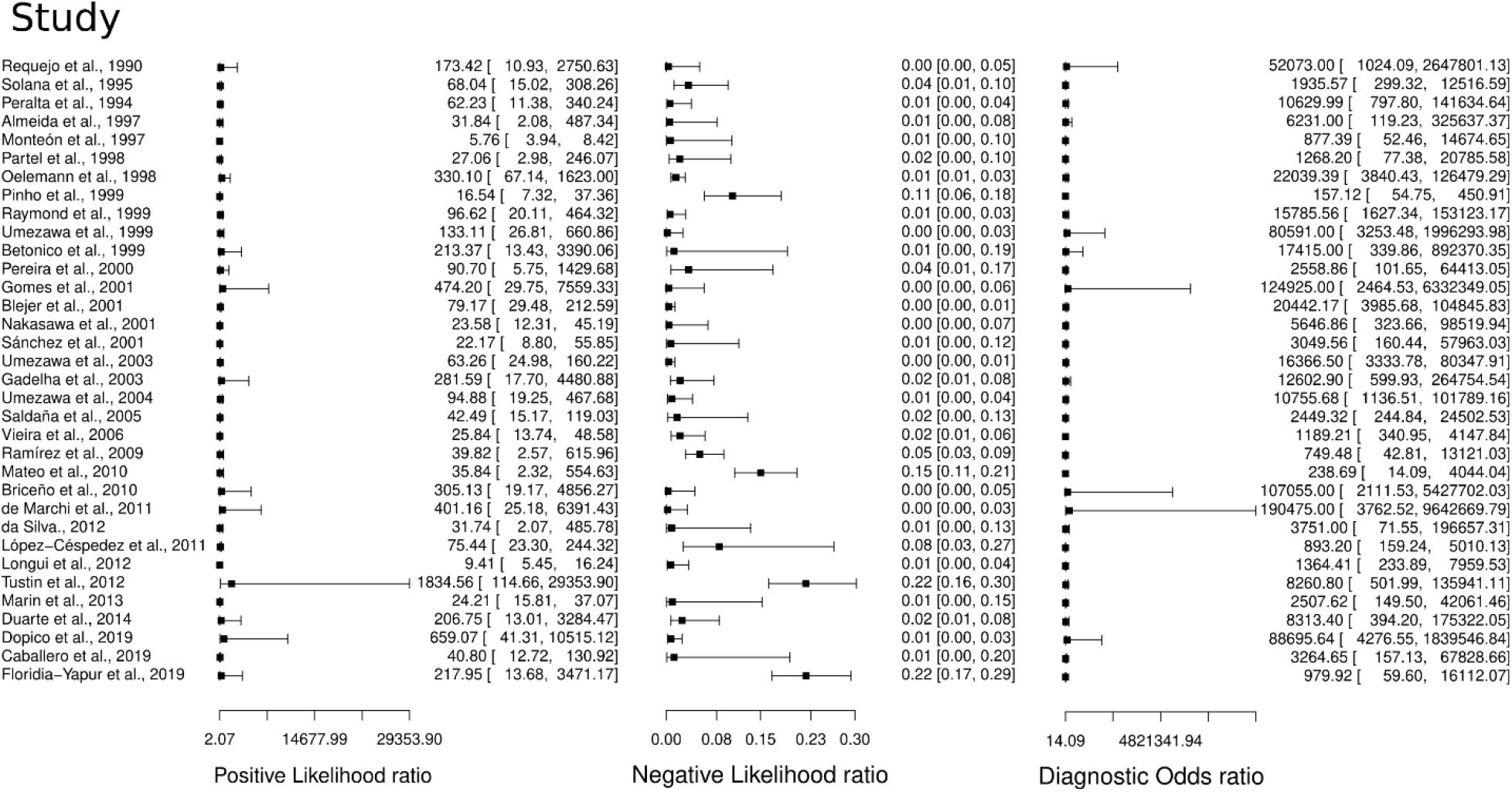
Study data and paired forest plot of the Positive Likelihood ratio, Negative likelihood ratio, and Diagnostic Odds ratio of Enzyme-linked immunosorbent assay (ELISA) in Chagas’s disease diagnosis. Data from each study are summarized. Positive Likelihood ratio, Negative likelihood ratio, and Diagnostic Odds ratio are reported with a mean (95% confidence limits). The Forest plot depicts the estimated sensitivity and specificity (black squares) and its 95% confidence limits (horizontal black line).

**Supplementary 2.**
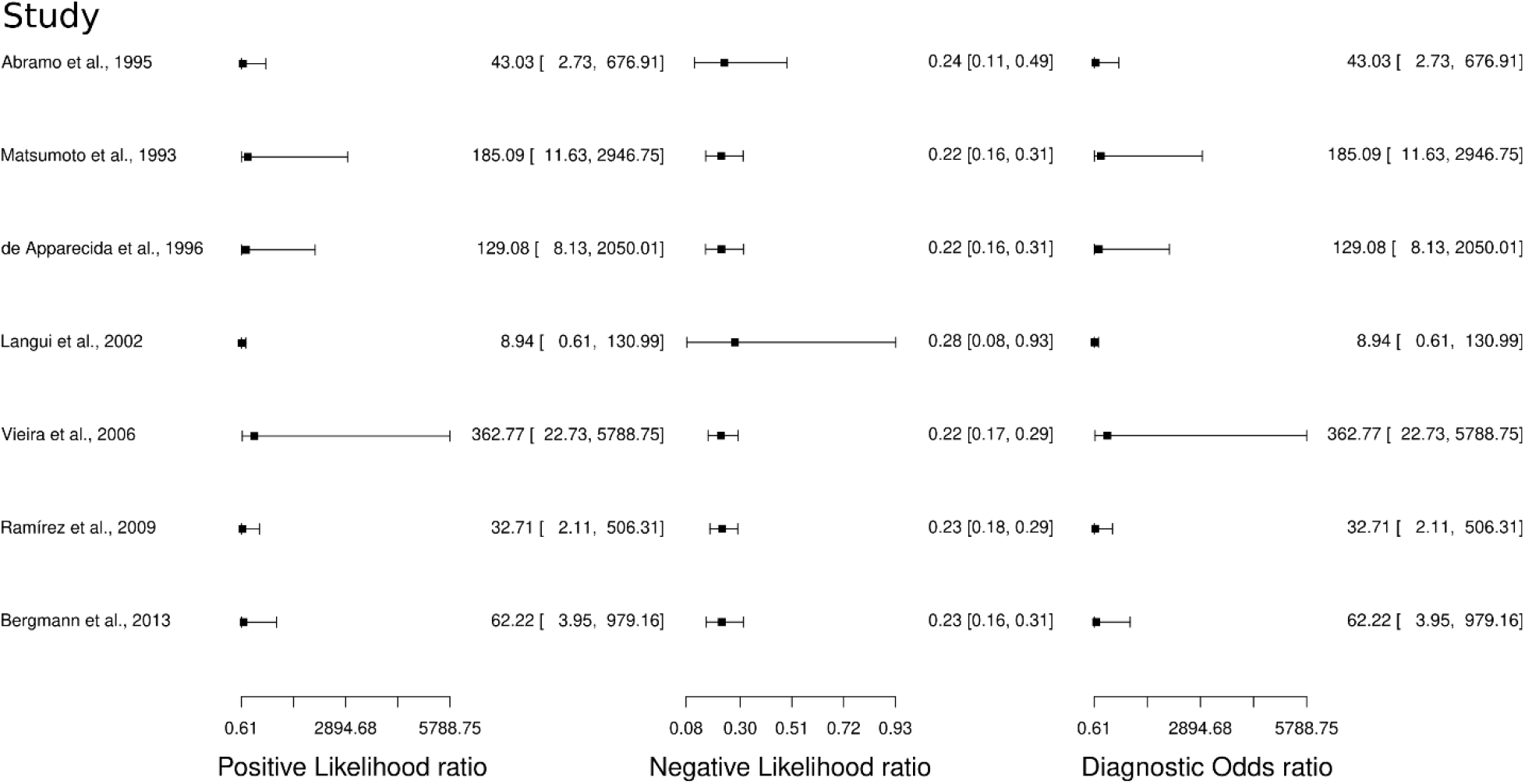
Study data and paired forest plot of the Positive Likelihood ratio, Negative likelihood ratio, and Diagnostic Odds ratio of Fluorescence antibody assay (FAT) in Chagas’s disease diagnosis. Data from each study are summarized. Positive Likelihood ratio, Negative likelihood ratio, and Diagnostic Odds ratio are reported with a mean (95% confidence limits). The Forest plot depicts the estimated sensitivity and specificity (black squares) and its 95% confidence limits (horizontal black line).

**Supplementary 3.**
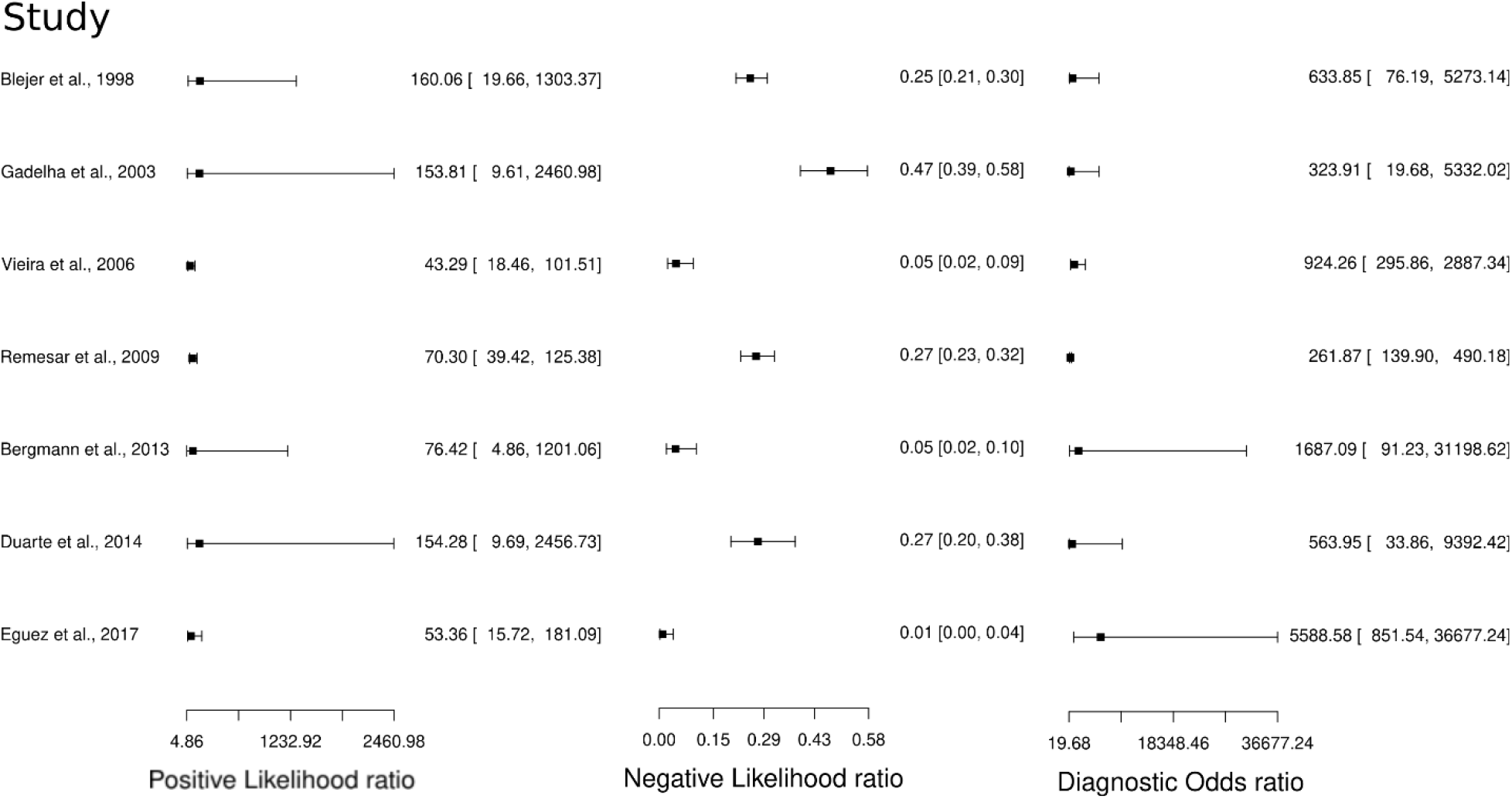
Study data and paired forest plot of the Positive Likelihood ratio, Negative likelihood ratio, and Diagnostic Odds ratio Hemagglutination test (HmT) in Chagas’s disease diagnosis. Data from each study are summarized. Positive Likelihood ratio, Negative likelihood ratio, and Diagnostic Odds ratio are reported with a mean (95% confidence limits). The Forest plot depicts the estimated sensitivity and specificity (black squares) and its 95% confidence limits (horizontal black line).

**Supplementary 4.**
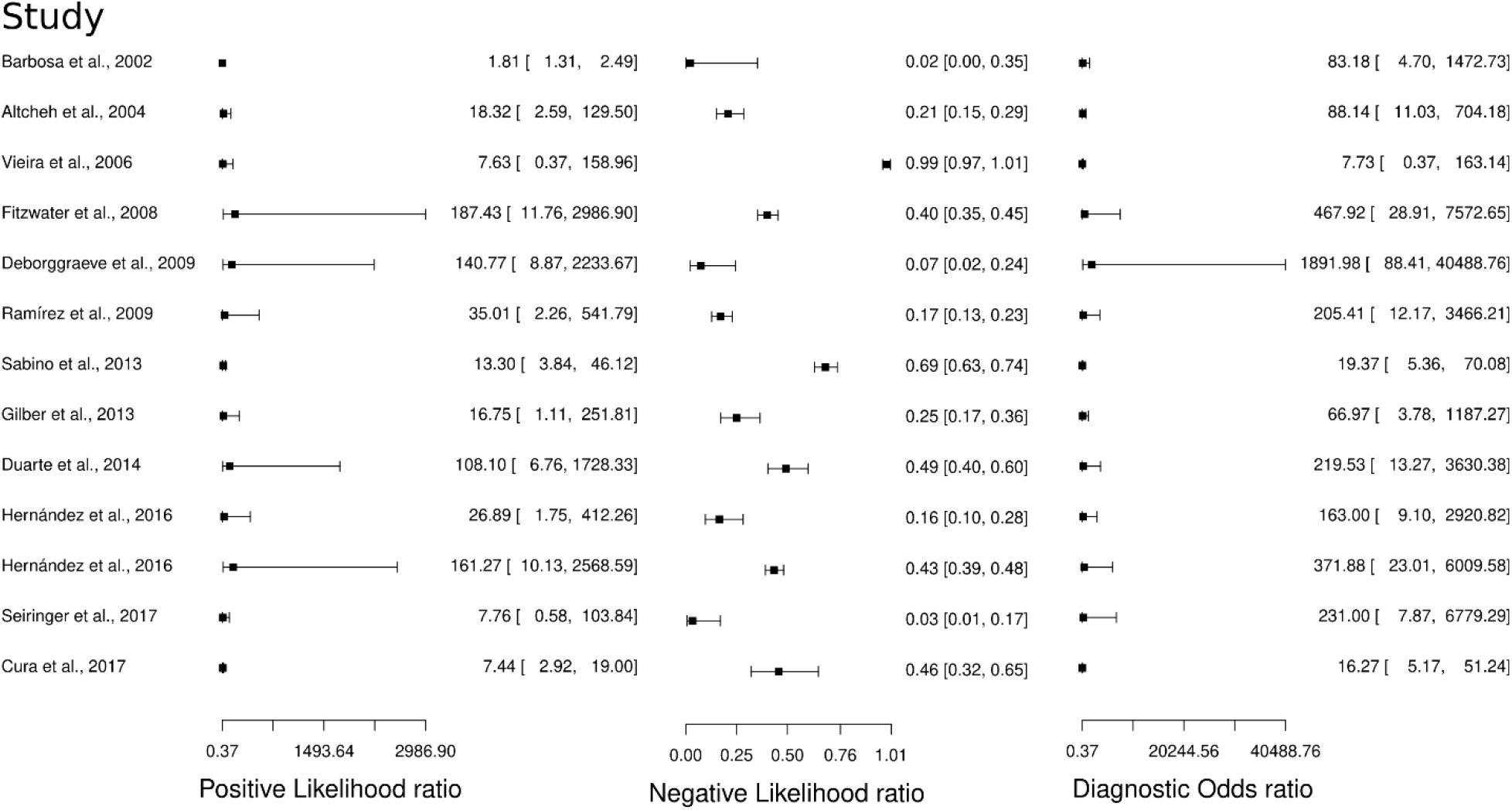
Study data and paired forest plot of the Positive Likelihood ratio, Negative likelihood ratio, and Diagnostic Odds ratio Polymerase chain reaction in Chagas’s disease diagnosis. Data from each study are summarized. Positive Likelihood ratio, Negative likelihood ratio, and Diagnostic Odds ratio are reported with a mean (95% confidence limits). The Forest plot depicts the estimated sensitivity and specificity (black squares) and its 95% confidence limits (horizontal black line).

**Supplementary 5.**
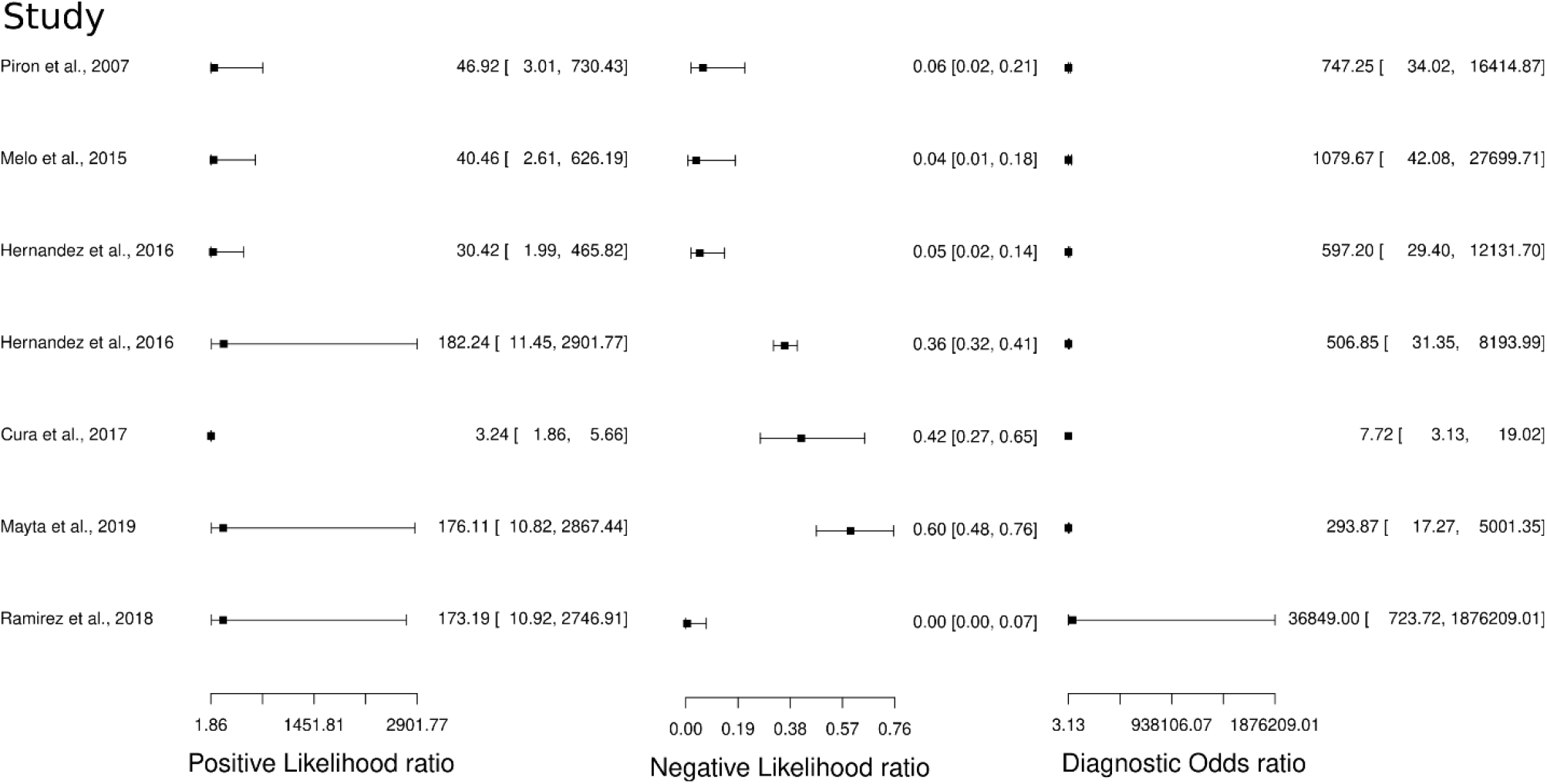
Study data and paired forest plot of the Positive Likelihood ratio, Negative likelihood ratio, and Diagnostic Odds ratio quantitative Polymerase chain reaction in Chagas’s disease diagnosis. Data from each study are summarized. Positive Likelihood ratio, Negative likelihood ratio, and Diagnostic Odds ratio are reported with a mean (95% confidence limits). The Forest plot depicts the estimated sensitivity and specificity (black squares) and its 95% confidence limits (horizontal black line).

